# Detection of SARS-CoV-2 specific IgA in the human milk of COVID-19 vaccinated, lactating health care workers

**DOI:** 10.1101/2021.04.02.21254642

**Authors:** Vivian Valcarce, Lauren S. Stafford, Josef Neu, Nicole Cacho, Leslie Parker, Martina Mueller, David J Burchfield, Nan Li, Joseph Larkin

## Abstract

**Importance:** In 2019, a deadly virus known as severe acute respiratory syndrome coronavirus 2 (SARS-CoV-2), responsible for COVID-19, emerged. In December 2020, two mRNA-based COVID-19 vaccines were approved for use in the United States (US) which provide immunity to those receiving the vaccine. Maternally derived antibodies are a key element of infants’ immunity. Certain vaccines given to pregnant and lactating mothers provide immunity to infants through transmission across the placenta, umbilical cord (IgG) and human milk (IgA). Human milk produced by mothers with a history of COVID-19 infection contains SARS-CoV-2 IgA and IgG.

**Objective:** To determine whether SARS-CoV-2 specific immunoglobulins are found in human milk after the COVID-19 vaccination, and to characterize the types of immunoglobulins present.

**Design, setting, and participants:** This is a prospective observational study conducted at Shands Hospital, University of Florida from December 2020 to March 2021. Twenty-two lactating healthcare workers who received the SARS-CoV-2 mRNA vaccine (Pfizer/BioNtech or Moderna) made up the sample group. Plasma and human milk were collected at three-time points (pre-vaccination, post-first vaccine dose, and post-second vaccine dose). SARS-CoV-2 specific IgA and IgG in human milk and in plasma were measured by ELISA. Maternal demographics was compiled.

**Exposures:** Pfizer/BioNtech or Moderna vaccination.

**Main outcome and measure:** Levels of SARS-CoV-2 IgA and IgG in human milk and plasma.

**Results:** We found significant secretion of SARS-CoV-2 specific IgA and IgG in human milk and plasma after SARS-CoV-2 vaccination.

**Conclusions and relevance:** Our results show that the mRNA-based COVID-19 vaccines induce SARS-CoV-2 specific IgA and IgG secretion in human milk. Further studies are needed to determine the duration of this immune response, its capacity to neutralize the COVID-19 virus, the transfer of passive immunity to breastfeeding infants, and the potential therapeutic use of human milk IgA to combat SARS-CoV-2 infections and COVID-19.

**KEY POINTS:** *Question:* Is there SARS-CoV-2 specific IgA in the human milk of lactating women after COVID-19 vaccination?

*Findings:* In this prospective observational study that included 22 lactating women, we found SARS-CoV-2 specific IgA in the human milk in response to the COVID-19 vaccination series. There is statistically significant secretion of SARS-CoV-2 IgA in human milk after mRNA COVID-19 vaccination series completion (p < 0.0001).

*Meaning:* Newborn immunologic defense is present but immature. SARS-CoV-2 IgA secreted in the human milk could potentially provide COVID-19 protection to nursing infants. These results could guide a strategy for SARS-CoV-2 vaccination among lactating women.

## INTRODUCTION

In 2019, severe acute respiratory syndrome coronavirus 2 (SARS-CoV-2) emerged, causing an outbreak of COVID-19 disease responsible for over 2.5 million deaths worldwide. In response, two mRNA vaccines, the BNT162b2 Pfizer/BioNTech and mRNA-1273 Moderna/NIH, emerged in record time and were approved for emergency use in the United States. Both vaccines contain mRNA encapsulated in a lipid nanoparticle, encoding SARS-CoV-2 Spike protein. Data indicate that two repeated parenteral injections are generally safe and induce strong protein-specific antibody responses as well as CD4+ and CD8+ T cell responses.^1^ Preliminary reports for the mRNA-based COVID-19 vaccines phase I/II trials showed significant SARS-CoV-2 IgG antibody response after vaccination series completion, similar to those observed in convalescent human serum samples of diagnosed COVID-19 individuals.^2,3^ Also, two doses of either SARS-CoV-2 mRNA-based vaccines were safe and provided 94-95% efficacy against symptomatic COVID-19 in persons 18 and older.^4,5^

Vaccinating pregnant or lactating mothers is a strategy to protect young infants from disease.^6^ In general, vaccines mediate protection through the induction of antigen-specific antibodies. Some vaccines, such as Rotavirus, oral Polio, and nasal Influenza, induce serum IgA and secretory IgA production.^7^ Vaccination against pertussis in the second/third trimester of pregnancy or immediately postpartum significantly increases the levels of anti-pertussis secretory IgA in human milk.^6^

As per the American Academy of Pediatrics report from February 2021, children represent up to 13% of the total cumulative cases of COVID-19, with up to 3% of those requiring hospitalization and up to 0.25% resulting in death.^8^ For these reasons, there is an urgent need to study human milk for SARS-CoV-2 specific antibody response to COVID-19 vaccination and, more importantly, its possible extended protection to breastfeeding infants.

In human milk, 90% of total immunoglobin is secretory IgA, along with 8% IgM and 2% IgG. Mother’s milk immunoglobulins delivered to infants during breastfeeding are crucial in shaping and modulating immature infants’ immune system and play an essential role in neutralization and agglutination processes, limiting the ability of pathogens to infect or persist.^9^

Although several studies demonstrate that milk produced by mothers, previously infected by COVID-19 is a source of neutralizing anti-SARS-CoV-2 IgA and IgG antibodies, ^10–12^ there is a paucity of data related to vaccination. Two studies demonstrating the presence of SARS-CoV-2 IgA and IgG in human milk following the COVID-19 vaccine are currently under peer review.^13,14^ This study’s primary purpose is to determine the presence of specific SARS-CoV-2 IgA in the human milk of lactating women after the COVID-19 vaccine administration. We hypothesize that the COVID-19 vaccine will elicit the production of SARS-CoV-2 IgA in human milk that could be passively transferred to breastfeeding infants.

## METHODS

### Study design

This University of Florida IRB-approved (protocol #202003255) study includes 22 lactating healthcare workers, after COVID-19 vaccine administration from December 2020 to March 2021. Please see supplemental methods for inclusion criteria, participants, and procedures.

### Sample collection and processing

Blood and human milk samples were collected at three-time points guided by the preliminary data from Pfizer/BioNtech COVID-19 vaccine^2^: pre-vaccination (TP1), 15-30 days post the first dose of vaccine (TP2), and 7-10 days post-second dose of vaccine (TP3). (See supplementary results)

### Statistical analysis

The focus of this analysis was change in titer levels over time and differences in changes between vaccine types.^15^ A mixed-effects repeated-measures modeling approach, which accommodates missing data, was used to model change in log (10)-transformed antibody titer for SARS-CoV-2 specific and total IgG/IgA in plasma and human milk over time (at pre-vaccination, post 1^st^ and post 2^nd^ dose). (Detailed Statistical analysis present in supplemental)

## RESULTS

Twenty-two lactating health care workers with no known history of COVID-19 infection were enrolled in the study. Of those, 21 completed the three-time sample collection for human milk: pre-vaccination (TP1), post 1^st^ vaccine (TP2), and post 2^nd^ vaccine (TP3). The study population consisted primarily of White, non-Hispanic women in their mid-30s working in the healthcare setting. Seven participants received the Moderna vaccine and 14 received the Pfizer/BioNTech vaccine (Table 1).

**Table 1.** Study participants characteristics (n= 21)^a^

| **Maternal characteristics (n=21)** | **N (%) or Mean ± Standard deviation** |
| --- | --- |
| Age (yr) | 34 ± 3.9 |
| Race |  |
| *White* | 20 (95) |
| *Asian* | 1 (5) |
| Ethnicity |  |
| *Non- Hispanic* | 18 (85) |
| *Hispanic* | 3 (15) |
| Profession |  |
| *Healthcare worker* | 19 (90) |
| Body mass index (kg/m<sup>2</sup>)<sup>b*</sup> | 24.8 ± 3.4 |
| *Normal/Healthy weight* | 11 (55) |
| *Overweight* | 7 (35) |
| *Obese* | 2 (10) |
| History of allergies* | 6 (30) |
| History of asthma* | 1 (5) |
| History of inadequate immune response to vaccine* | 2 (10) |
| Family history of Cancer* | 13 (65) |
| Family history of autoimmune disorder* | 1 (5) |
| Antibiotic use last 6 mo* | 5 (25) |
| Parity (no.) | 1.9 ± 0.9 |
| Time postpartum (mo) | 6.8 ± 4.8 |
| Infant sex |  |
| *Female* | 12 (57) |
| *Male* | 9 (43) |
| Tandem breastfeeding | 1 (5) |
| Vaccine brand |  |
| *Moderna* | 7 (33.3) |
| *Pfeizer* | 14 (66.7) |
| Developed decreased human milk supply after COVID vaccine* | 3 (15) |

<sup>a</sup>Categorical data are given as number of participants and, in parentheses, percentage of total. Continuous data are provided as means ± standard deviations. <sup>b</sup>Definitions put forth by the U.S. Centers for disease Control and Prevention were used for body mass index categories. \*, missing data from 1 subject.

### 1. SARS-CoV-2 IgA is present in plasma and secreted in human milk after COVID-19 vaccination

SARS-CoV-2 IgA concentration in human milk was grouped and compared between the 3 time points. IgA statistically significantly increased from TP1 to TP2 (p < 0.0007) and from TP1 to TP3 (p < 0.0001) (Figure 1) in the overall group of participants. Eighty-five percent of participants had a positive result for SARS-CoV-2 IgA after vaccination series completion based on the established cutoff value (mean + 2SD of pre-vaccination SARS-CoV-2 IgA concentration values) (Table 2); this result coincides with the IgA response to the natural infection (76-80%).^10–12^

**Figure 1.**
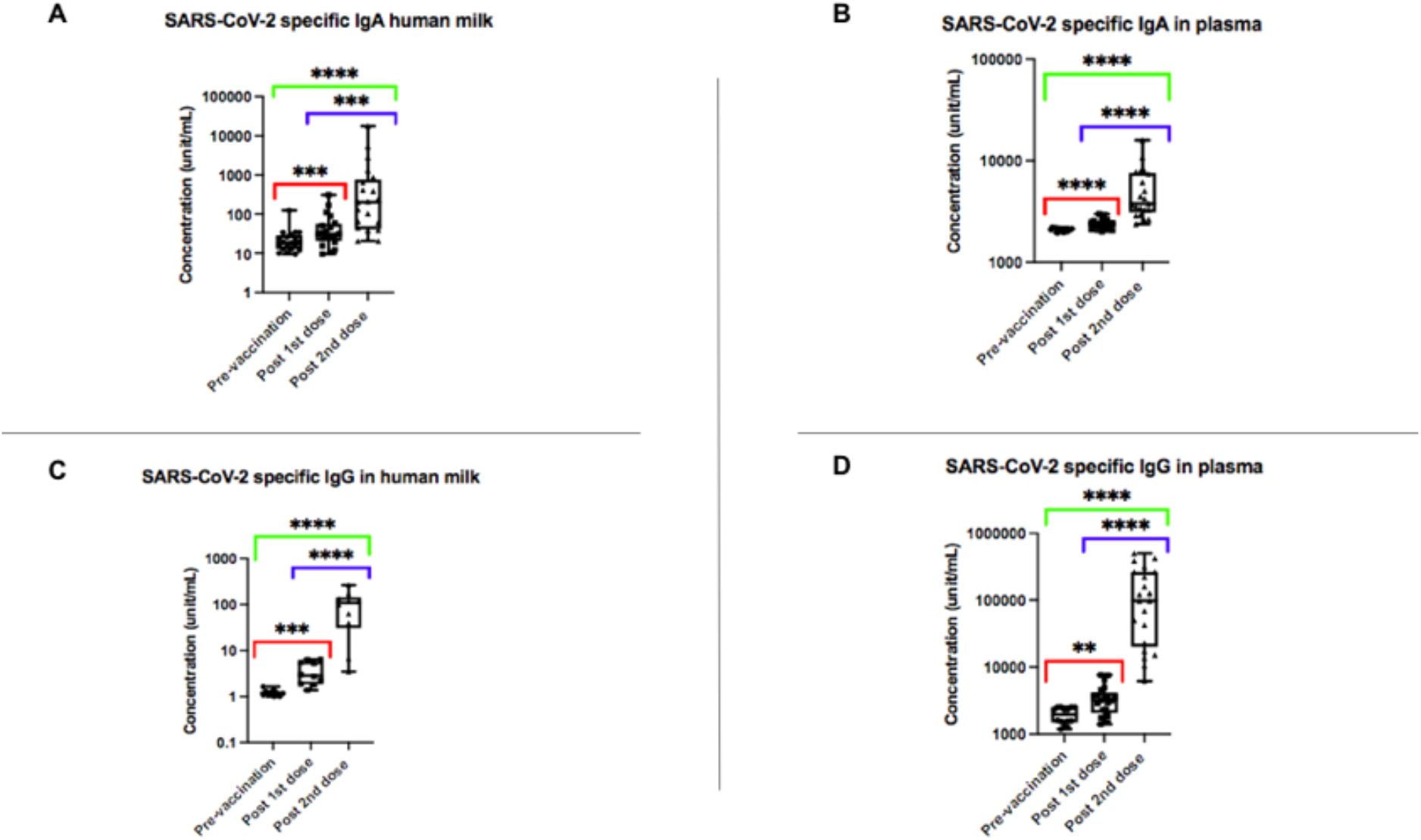
Box and whisker plots of SARS-CoV-2 specific antibodies (IgA and IgG) in human milk and plasma pre-vaccination, post 1^st^ dose of vaccine, and post 2^nd^ dose of vaccine measured as unit/mL (a) IgA in human milk (b) IgA in plasma (c) IgG in human milk and (d) IgG in plasma. All concentrations are shown as log(10). ** p < 0.01; *** p < 0.001; **** p < 0.0001.

**Table 2.** SARS-CoV-2 antibody positivity post COVID-19 vaccination

Eighty five percent of human milk samples were positive for SARS-CoV-2 IgA based on the established cut off value (mean + 2SD of pre-vaccination SARS-CoV-2 IgA concentration values). All human milk and plasma samples tested for SARS-CoV-2 IgG were positive.
| Sample ID | Positive reactivity for SARS-CoV-2 specific antibodies after COVID-19 vaccination |  |  |  |
| --- | --- | --- | --- | --- |
|  | IgA Human milk | IgG Human milk | IgA Plasma | IgG Plasma |
| C-011224 | + | + | + | + |
| C-021230 | - | + | - | + |
| C-030101 | + | n/a | + | + |
| C-040103 | + | n/a | + | + |
| C-060106 | + | n/a | - | + |
| C-070106 | + | n/a | + | + |
| C-080106 | + | n/a | + | + |
| C-090107 | + | n/a | + | + |
| C-100108 | + | n/a | + | + |
| C-110108 | - | + | - | + |
| C-120110 | + | + | + | + |
| C-130111 | + | n/a | + | + |
| C-140112 | + | n/a | + | + |
| C-150113 | + | n/a | + | + |
| C-160121 | + | n/a | + | + |
| C-170121 | + | + | + | + |
| C-18 0127 | - | + | + | + |
| C-190204 | + | + | + | + |
| C-200204 | + | + | + | + |
| C-210209 | + | + | + | + |
| C-220217 | + | + | + | + |

SARS-CoV-2 IgA in plasma also significantly increased 7-10 days after vaccination series completion and was positively correlated with SARS-CoV-2 IgA in human milk (p < 0.0001) (Supplementary Table 2).

### 2. SARS-CoV-2 IgG is secreted in human milk after COVID-19 vaccination

We tested human milk for SARS-CoV-2 IgG at the three-time points in 10 participants. One hundred percent of the human milk samples were found to be positive for SARS-CoV-2 IgG by TP3 based on the established cutoff value (Table 2). There was a statistically significant increase of IgG from TP1 to TP2 (p < 0.0006) and TP1 to TP3 (p < 0.0001) (Figure 1) (Supplementary Table 2). The concentration of SARS-CoV-2 IgA was higher than those of IgG in the human milk, with a predominance of SARS-CoV-2 IgA, as found in natural infection.^10–12^

### 3. Plasma samples are positive for SARS-CoV-2 IgG after COVID-19 vaccination

As expected, based on the preliminary reports for Pfizer/BioNTech and Moderna vaccines,^2,3^ SARS-CoV-2 IgG is detected in plasma after the first and second dose of the mRNA vaccines with a peak level 7-10 days after the second vaccine. We also found a positive correlation between plasma and breastmilk IgG levels (p = 0.043). (Figure 1) (Supplementary table 3) Levels of total IgA in human milk and Total IgG in plasma, were indistinguishable at all three-time points, as anticipated. (Supplementary Figure 3).

### 4. Similar antibody response to Pfizer/BioNtech and Moderna vaccines was detected

Both mRNA vaccines generated statistically significant SARS-CoV-2 specific IgA and IgG in breastmilk and plasma by TP3. There is a significantly higher mean of SARS-CoV-2 IgG in plasma at TP3 in Pfizer vs. Moderna vaccinated mothers (p = 0.005). (Supplementary Table 2).

## DISCUSSION

Our results show that the mRNA-based COVID-19 vaccines, Pfizer/BioNtech and Moderna, induce SARS-CoV-2 IgA and IgG secretion in human milk, with statistically significant increases from TP1 to TP3 (p < 0.0001 for IgA and p = 0.0002 for IgG). There is a predominant IgA response in human milk, as expected based on human milk antibody composition, and similar to the natural infection results.^10–12^ However, these results contrast with recent studies presented at medRxiv for peer review.^13,14^ The peak of SARS-CoV-2 IgA and IgG in human milk and plasma occurs 7-10 days after receiving the second dose of the COVID-19 vaccine. The peak of vaccine-induced antibody titers is but one part of antibody-mediated protection. The quality of such antibodies (e.g., their avidity, specificity, or neutralizing capacity) also contribute to efficacy.^7^ Further functional studies are needed to determine antibody neutralization capability and clinical relevance.

The samples analyzed represent only one time point of what is likely a dynamic immune response. Samples collected in the first 2 weeks after the second vaccine administration may not reflect the maximal immune response. Long-term follow-up is needed to understand the kinetics of this response, particularly the longevity of antibody presence.

Our data show higher SARS-CoV-2 IgG among mothers who received the Pfizer vaccine. However, statistical significance does not imply clinical relevance, and these results indicate that further, more extensive studies need to be conducted to confirm any findings.

There was not a statistically significant relationship between maternal demographics and the level of SARS-CoV-2 IgA in human milk (supplementary Table 3). Interestingly, participant C-190204 showed the highest concentration of SARS-CoV-2 IgA in the human milk (12 times higher than the mean of IgA post-vaccination) and was the only participant tandem breastfeeding her two children. Previous findings show a strong positive correlation between secretory IgA concentrations and lactation duration.^9^ Future studies, including twins, would be of interest, and a larger sample size with a broader demographic distribution is needed to generalize these results.

These novel results suggest the potential transfer of protective antibodies to nursing infants after maternal COVID-19 vaccination and may show a promising influence in vaccination strategy for lactating mothers.

## Data Availability

All data pertaining to this manuscript is contained within the manuscript

## FUNDING

Children’s Miracle Network

---

**Supplemental Table 2.** Least squares means and differences in least squares means with standard errors (and 95% confidence intervals) from longitudinal models investigating change in log(10) transformed antibody titers in plasma and breast milk over time, overall and by vaccine type.

| Log(10) transformed IgAs/IgGs overall | Pre-vaccination | Post 1<sup>st</sup> dose | Post 2<sup>nd</sup> dose | Pre-vaccination to post 1<sup>st</sup> dose | p-value | Pre-vaccination to post 2<sup>nd</sup> dose | p-value | Post 1<sup>st</sup> dose to post 2<sup>nd</sup> dose | p-value |
| --- | --- | --- | --- | --- | --- | --- | --- | --- | --- |
| SARS-CoV-2 specific IgA in breast milk N=21 | 1.41 ± 0.12 | 1.65 ± 0.13 | 2.34 ± 0.18 | -0.24 ± 0.06<br>(-0.37; -0.12) | 0.0007 | -0.93 ± 0.19<br>(-1.32; -0.53) | <0.0001 | -0.68 ± 0.17<br>(-1.04; -0.32) | 0.0008 |
| SARS-CoV-2 specific IgG in breast milk N=10 | 0.08 ± 0.02 | 0.50 ± 0.08 | 1.77 ± 0.20 | -0.42 ± 0.08<br>(-0.60; -0.23) | 0.0006 | -1.69 ± 0.19<br>(-2.12; -1.25) | <0.0001 | -1.27 ± 0.16<br>(-1.62; -0.92) | <0.0001 |
| SARS-CoV-2 specific IgA in plasma N=21 | 3.23 ± 0.01 | 3.38 ± 0.01 | 3.66 ± 0.05 | -0.05 ± 0.01<br>(-0.08; -0.03) | <0.0001 | -0.34 ± 0.05<br>(-0.44; -0.24) | <0.0001 | -0.29 ± 0.05<br>(-0.38; -0.19) | <0.0001 |
| SARS-CoV-2 specific IgG in plasma N=21 | 3.27 ± 0.04 | 3.49 ± 0.05 | 4.90 ± 0.13 | -0.23 ± 0.05<br>(-0.34; -0.11) | 0.0008 | -1.61 ± 0.13<br>(-1.90; -1.36) | <0.0001 | -1.41 ± 0.10<br>(-1.62; -1.19) | <0.0001 |
| Total IgA in breast milk N=8 | 6.39 ± 0.05 | 6.41 ± 0.06 | 6.37 ± 0.08 | -0.02 ± 0.04<br>(-0.12; 0.08) | 0.652 | 0.02 ± 0.04<br>(-0.07; 0.11) | 0.583 | 0.04 ± 0.05<br>(-0.08; 0.17) | 0.446 |
| Total IgG in plasma N=15 | 6.96 ± 0.05 | 6.81 ± 0.06 | 6.96 ± 0.05 | 0.14 ± 0.07<br>(-0.02; 0.31) | 0.076 | -0.003 ± 0.05<br>(-0.11; 0.11) | 0.947 | -0.15 ± 0.09<br>(-0.34; 0.05) | 0.127 |
| Log(10) transformed IgAs/IgGs by vaccine type* | Pre-vaccination | Post 1<sup>st</sup> dose | Post 2<sup>nd</sup> dose | Pre-vaccination to post 1<sup>st</sup> dose | p-value | Pre-vaccination to post 2<sup>nd</sup> dose | p-value | Post 1<sup>st</sup> dose to post 2<sup>nd</sup> dose | p-value |
| SARS-CoV-2 specific IgA in breast milk N=21<br>M: (n=7)<br>P: (n=14) | 1.52 ± 0.21<br>1.35 ± 0.15<br>p=0.508 | 1.71 ± 0.22<br>1.62 ± 0.16<br>p=0.756 | 2.60 ± 0.31<br>2.20 ± 0.22<br>p=0.304 | -0.19 ± 0.11<br>-0.27± 0.08<br>p=0.517 | 0.096<br>0.002 | -1.08 ± 0.33<br>-0.85± 0.24<br>p=0.584 | 0.004<br>0.002 | -0.89 ± 0.30<br>-0.58 ± 0.21<br>p=0.405 | 0.008<br>0.014 |
| SARS-CoV-2 specific IgA in plasma N=21<br>M: (n=7)<br>P: (n=14) | 3.32 ± 0.01<br>3.33 ± 0.01<br>p=0.574 | 3.36 ± 0.02<br>3.38 ± 0.01<br>p=0.241 | 3.72 ± 0.09<br>3.64 ± 0.06<br>p=0.443 | -0.04 ± 0.02<br>-0.06 ± 0.01<br>p=0.266 | 0.052<br>0.0003 | -0.40 ± 0.09<br>-0.31 ± 0.06<br>p=0.925 | 0.0002<br><0.0001 | -0.36 ± 0.08<br>-0.25 ± 0.06<br>p=0.276 | 0.0002<br>0.0002 |
| SARS-CoV-2 specific IgG in plasma N=21<br>M: (n=7)<br>P: (n=14) | 3.27 ± 0.07<br>3.27 ± 0.04<br>p=0.958 | 3.45 ± 0.08<br>3.52 ± 0.06<br>p=0.535 | 4.42 ± 0.18<br>5.13 ± 0.13<br>p=0.005 | -0.18 ± 0.10<br>-0.25 ± 0.07<br>p=0.471 | 0.086<br>0.003 | -1.16 ± 0.19<br>-1.87 ± 0.13<br>p=0.001 | <0.0001<br><0.0001 | -0.97 ± 0.14<br>-1.62 ± 0.10<br>p=0.001 | <0.0001<br><0.0001 |

\*M=Moderna; P=Pfizer; p=p-value for difference between M+P with Bonferroni-adjusted level of significance alpha: 0.0006

**Supplemental Table 3.** Beta coefficients with standard errors and 95% confidence intervals from longitudinal models investigating the relationship of log(10)-transformed SARS-CoV-2 specific IgA (dependent variable)/IgG in breastmilk with that in plasma as well as the relationship of log(10)-transformed SARS-CoV-2 specific IgA in breastmilk with maternal age, race, ethnicity, allergies, antibiotic use, BMI, and prior medical history over time.

|  | Beta | Standard error | 95% CI | p-value |
| --- | --- | --- | --- | --- |
| *Log-transformed SARS-CoV-2 specific IgA in breast milk vs plasma* | 2.76 | 0.52 | 1.69; 3.82 | <0.0001 |
| *Log-transformed SARS-CoV-2 specific IgG in breast milk vs plasma* | 0.44 | 0.12 | 0.03; 0.86 | 0.043 |
| **Log-transformed SARS-CoV-2 specific IgA** |  |  |  |  |
| *Maternal age* | 0.01 | 0.03 | -0.06; 0.08 | 0.815 |
| *Maternal race (Reference group: White)* | -0.43 | 0.60 | -1.68; 0.82 | 0.484 |
| *Ethnicity (Reference group: Not Hispanic)* | -0.07 | 0.33 | -0.77; 0.62 | 0.827 |
| *Allergies (Reference group: Yes))* | 0.33 | 0.29 | -0.29; 0.94 | 0.282 |
| *Infant age /length breastfeeding* | 0.04 | 0.03 | -0.02; 0.09 | 0.146 |
| *Used antibiotic in past 6 months (Reference group: Yes))* | -0.04 | 0.32 | -0.73; 0.64 | 0.888 |
| *History of "Other" (Reference group: No)* | -0.18 | 0.46 | -1.16; 0.80 | 0.705 |
| *History of asthma (Reference group: No)* | -0.26 | 0.64 | -1.61; 1.08 | 0.685 |
| *Family history*<br>*Autoimmune disease (Reference group: None)*<br>*Cancer (Reference group: None)* | -0.32<br>0.12 | 0.70<br>0.33 | -1.79;<br>1.16<br>-0.59;<br>0.83 | 0.782 |
| *History of not mounting an appropriate response to a vaccine* | 0.53 | 0.44 | -0.40; 1.47 | 0.246 |
| *Maternal BMI* | 0.002 | 0.01 | -0.01; 0.02 | 0.722 |

**Supplemental Figure 2.**
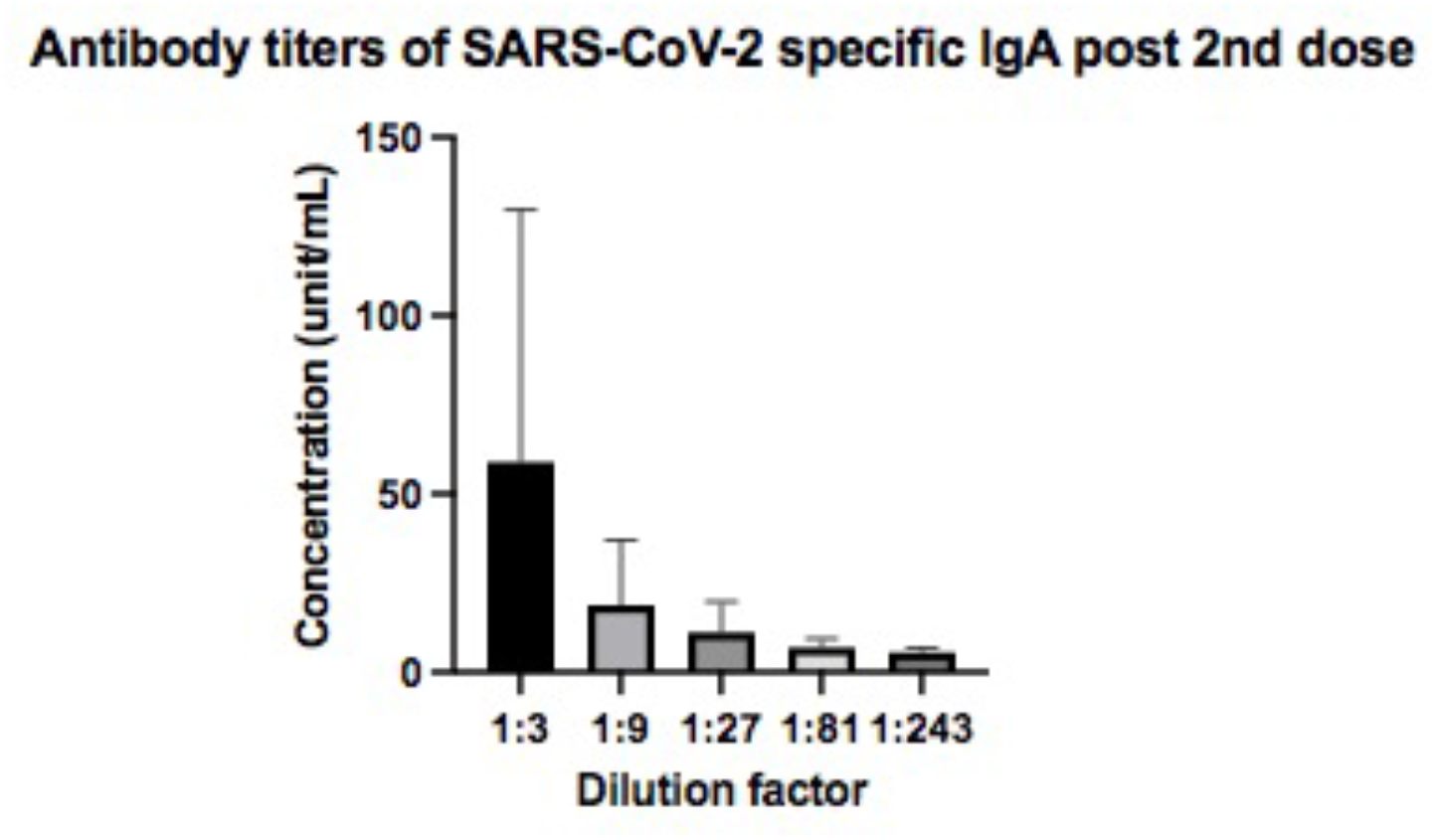
Antibody titers of SARS-CoV-2 specific IgA in human milk post second dose of vaccine

**Supplemental Figure 3.**
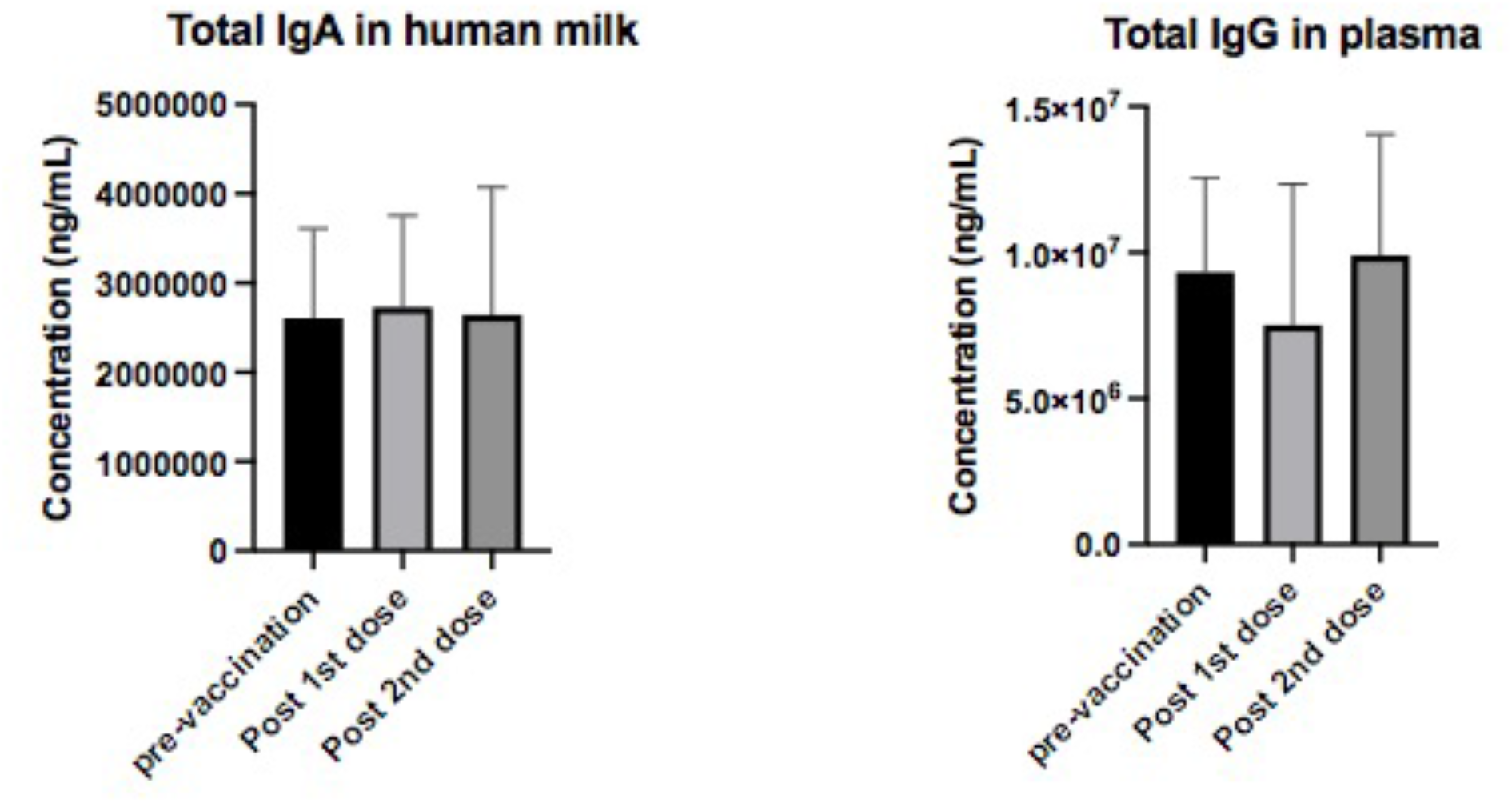
Total antibody concentration in human milk (IgA) (n=8) and in plasma (IgG) (n=15)

## Notes

### Competing Interest Statement

The authors have declared no competing interest.

### Funding Statement

The work presented in this manuscript was funded by the Childrens Miracle Network. No authors received payment or services for any aspect of the published work

### Author Declarations

Dr. Margaret Hamer of the University of Florida Institutional Review Board (UF IRB) led the evaluation of protocol. The UF IRB granted ethical approval our study. This prospective observational IRB-approved (protocol 202003255) study included 22 lactating healthcare workers, each one followed for up to 42 days after the first COVID19 vaccine administration from December 2020 to March 2021. Inclusion criteria were lactating women with no known history of COVID19 infection; greater than 18 years old, able to provide informed consent, and receiving the COVID19 vaccine. Seven participants had no prevaccination blood sample collected due to late enrollment. One subject could not complete sample collection and thus was not included in our analysis.

